# Characteristics of Patients that Substitute Medical Cannabis for Alcohol

**DOI:** 10.1101/19003798

**Authors:** Assad Hayat, Brian J. Piper

## Abstract

**Aims:** A substitution effect occurs when patients substitute Medical Cannabis (MC) for another drug. Over three-quarters (76.7%) of New England dispensary members reported reducing their use of opioids and two-fifths (42.0%) decreased their use of alcohol after starting MC (Piper et al. 2017). The objective of this exploratory study was to identify any factors which differentiate alcohol substituters from those that do not modify their alcohol use after starting MC (non-substituters).

**Methods:** Among dispensary patients (N=1,477), over two-thirds with chronic pain, that completed an online survey, 7.4% indicated that they regularly consumed alcohol. Comparisons were made to identify any demographic or health history characteristics which differentiated alcohol substituters (N=47) from non-substituters (N=65). Respondents selected from among a list of 37 diseases and health conditions (e.g. diabetes, sleep disorders) and the total number was calculated.

**Results:** Substituters and non-substituters were indistinguishable in terms of sex, age, or prior drug history. Substituters were significantly more likely to be employed (68.1%) than non-substituters (51.1%). Substituters also reported having significantly more health conditions and diseases (3.3±2.0) than non-substituters (2.4±1.4).

**Conclusions:** This small study offers some insights into the profile of patients whose self-reported alcohol intake decreased following initiation of MC. Alcohol substituters had more other health conditions but also were more likely to be employed which may indicate that they fit a social drinker profile. Additional prospective or controlled research into the alcohol substitution effect following MC with a sample with more advanced alcohol misuse may be warranted.

**Short summary:** A substitution effect with medical cannabis replacing prescription opioids has been reported but less is known for alcohol. This study evaluated characteristics which might differentiate alcohol substituters (N=47) from non-substituters (N=65) among dispensary members. Substituters were significantly more likely to be employed and have more health conditions than non-substituters.

---

There has been a substantial amount of clinical and epidemiological research of the potential therapeutic benefits of medical cannabis (MC), particularly in the past two decades (National Academy of Sciences, 2017). In the US, MC is currently legal in twenty-nine states and also in Washington DC, Guam, and Puerto Rico. Further, nineteen states condone low tertrahydrocannabinol (THC) / high cannabidiol (CBD) products (National Conference of State Legislatures, 2017) and eight states have legalized cannabis for recreational use. The regulations for the use of MC are state specific. States have approved the use of MC for a variety of diseases and disorders including pain, spasticity associated with multiple sclerosis, nausea, post-traumatic stress disorder (PTSD), cancer, epilepsy, cachexia, glaucoma, HIV/AIDS, and degenerative neurological diseases (National Organization for the Reform of Marijuana Laws, 2017) with chronic pain being the most common condition (Reiman et al. 2017).

The US is in the midst of an opioid epidemic involving both commercial pharmaceutical (e.g. oxycodone, hydrocodone) and illicitly manufactured (e.g. heroin, fentanyl) drugs (Piper et al. 2016; Wright et al. 2014). As a result, there is tremendous interest in whether MC users decrease their use of opioid medications. Three lines of evidence are congruent with a MC induced opioid substitution effect. First, there have been several well-powered (N > 1,000) retrospective surveys from California (Reiman et al. 2017), New England (Piper et al. 2017a), and from across North America (Corroon et al. 2017) which have identified a substitution effect. However, although results from these samples collectively indicate that this opioid substitution effect is a robust phenomenon, self-reported data from voluntary (i.e. self-selected) participants regarding this controversial drug should be viewed with great caution. Second, US states with MC had a 25% lower opioid overdose mortality rate relative to states without MC (Bachhuber et al. 2014). Third, states with MC have fewer opioid expenditures for Medicare and Medicaid (Bradford & Bradford, 2016, 2017).

Although the opioid substitution effect is moderately well-established (although see Caputi, 2017), less is known about whether MC patients decrease their use of alcohol (Subbaraman, 2014). Excessive alcohol consumption exerts a significant burden on health care systems (Mehta, 2017). Two large (N > 35,000 each) nationally representative surveys found that over one out of every eight (12.7%) adults met criteria for an Alcohol Use Disorder (AUD, Grant, Chou, Saha, et al. 2017). Approximately 65,000 people (> 80% male) in the US die from alcohol related causes each year (Rehm, Dawson, Frick, et al. 2014). Currently available interventions for AUDs have modest efficacies and there are limited tools available to identify the subset of patients that may benefit. A meta-analysis of Alcoholics Anonymous (AA) and other 12-step programs revealed that the available evidence did not demonstrate effectiveness of AA to achieve abstinence, reduce alcohol use, or improve participation in treatment (Ferri, Amato, & Davoli, 2006). Further, the AA philosophy is very concerned about recovering alcoholics becoming addicted to prescription drugs or switching to marijuana (Alcoholics Anonymous, 2011). Although there are Food and Drug Administration approved pharmacotherapies for alcoholism, there is a need for agents that are both well-tolerated and efficacious. Randomized controlled trials with blind designs showed no efficacy of disulfiram relative to controls (Skinner et al. 2014). The number needed to treat (NNT) with acamprosate such that one person would show decreased likelihood of returning to drinking was nine (Rosner et al. 2011) or twelve (Jonas et al. 2014). The NNT with the opioid antagonist naltrexone was 20 (Jonas et al. 2014).

There is evidence using a variety of research designs which has evaluated whether cannabinoids may have a therapeutic role for alcohol misuse. Pyrahexyl, a synthetic homologue of THC, was administered to seventy alcoholics. The vast majority (84%) were classified as showing an alleviation of symptoms including calmness and decreased irritability during alcohol withdrawal (Thompson & Proctor, 1953). Further, Mrs. A, a 49 year-old woman who struggled with alcoholism, consumed less alcohol on weekends when she smoked marijuana socially. Her psychiatrist instructed her to switch to daily marijuana use which resulted in improved interpersonal outcomes (Mikuriya, 1970a, 1970b). Subsequently, ninety-two patients, two-thirds with alcoholism or cirrhosis of the liver, used cannabis to decrease their alcohol. Cessation of cannabis was also associated with a return to drinking (Mikuriya, 2004). A survey of MC dispensary members in northern California identified a general substitution effect for alcohol, illegal, and prescription drugs (Reiman, 2006). The rationale for MC substitution was fewer side effects and improved symptom management (Reiman, 2009, Lucas et al. 2015). Among New England dispensary patients that also regularly consumed alcohol, two-fifths indicated that they reduced their alcohol consumption (Piper et al. 2017a). In addition to clinical research (Subbaraman, 2014), there is also a preclinical evidence base showing that manipulating the endocannabinoid system impacts ethanol consumption (Sloan, Gowin, Ramchandani, et al. 2017; Viudez-Martínez A, García-Gutiérrez MS, Navarrón et al. 2017). Therefore, the objective of this exploratory study was to further characterize this substitution effect and identify any factors which differentiated alcohol substituters from non-substituters.

## METHODS

### Participants

Participants of this study were members of MC dispensaries located in New England. Additional information about the participants is available elsewhere (Piper, et al. 2017a, 2017b, Piper, 2018).

### Procedure

Participants received an email from their dispensary inviting them to complete surveys hosted on Survey Monkey. There were three slightly tailored survey versions (Vermont, Maine, Rhode Island) for a subset of items as different states have different MC laws. Responses were exported to spreadsheets and consolidated into a single file. The survey (Supplementary Appendix) used skip logic to tailor items based on prior responses. An affirmative response to “Do you regularly consume alcohol?” would result in additional items including “Have you noticed a change in your alcohol consumption since you started medical cannabis?” with options of “Yes, I need to drink a lot more”, “Yes, I need to drink slightly more”, “No change”, “Yes, I need to drink slightly less”, or “Yes, I need to drink a lot less”. Among all participants (N = 1,477), only those that reported regular alcohol use (N = 112, 50.4% male) are the topic of this investigation. Among this subset, the “No change” (N = 65) were classified as non-substituters while the “a lot less” (N = 27) and “slightly less” (N = 28) were collapsed into a substituter group (N = 55, note that no respondents reported an increase in alcohol consumption). The survey included a required informed consent, demographic information, MC use, and items about history of use of recreational drugs. Respondents that identified as employed full-time, part-time, or as students were classified as employed. The total number selections in response to “Which of the following diseases or conditions have you been diagnosed by a medical professional (physician, PA, psychologist) as having?” with 37 options (e.g. diabetes, high blood pressure, chronic pain, sleep disorder) was calculated as an index of overall health. This study, including the informed consent, was approved by the IRB of Maine Medical Center. Further information on the procedure was previously reported (Piper et al. 2017a, 2017b, Piper, 2018).

### Statistical Analysis

Differences between substituters and non-substituters were assessed using either an independent samples t-test or chi–square with a *p* < .05 considered significant. Variability was reported as the standard deviation in the text and the standard error of the mean in figures. Figures were prepared with GraphPad Prism, v. 6.07 (La Jolla, CA).

## RESULTS

Over three-fifths of respondents were from Vermont (62.5%), followed by Rhode Island (26.8%), and Maine (5.4%). Table 1 shows that substituters and non-substituters were equivalent in terms of age, sex, and ethnicity. Responses to “How would you describe your use of cannabis?” on a continuum, with 10% increments, from 100% recreational/0% medical to 0% recreational/100% medical, were on the medical end of the spectrum. The age of first smoking, drinking alcohol, and using marijuana recreationally were not statistically different between substituters and non-substituters. Ratings of “How effective is medical cannabis in treating your symptoms or condition(s)?” with options of 0% (no relief) to 100% (complete relief) were high but did not differ based on alcohol substitution.

**Table 1.** Comparison of patients that did (substituters) or did not (non-substituters) decrease their use of alcohol after starting medical cannabis. ddefined in results section, SD: Standard Deviation

|  | Substituters<br>(N = 47) | Non-Substituters<br>(N = 65) |
| --- | --- | --- |
| Age ( +SD) | 50.6 (12.1) | 52.6 (14.0) |
| Female (%) | 42.6% | 54.7% |
| White (%) | 91.8% | 100.0% |
| Body Mass Index (SD) | 26.8 (4.2) | 26.8 (4.7) |
| Prior recreational marijuana use (%) | 95.7% | 89.2% |
| Recreational vs Medical Used (SD) | 81.3 (20.4) | 82.2 (19.2) |
| Age first smoked cigarette (SD) | 14.5 (3.1) | 15.0 (2.8) |
| Age first drank alcohol (SD) | 15.3 (2.9) | 16.1 (2.8) |
| Age first used marijuana (SD) | 15.7 (2.2) | 16.2 (1.8) |
| Effective is MC in treating symptomsd (SD) | 74.7 (17.3) | 73.1 (18.2) |
| Route of administration (% joint) | 42.6% | 40.0% |
| Indica strain preference (%) | 38.6% | 47.7% |
| Amount (\$) spent on MC each month (SD) | 174.2 (138.6) | 134.3 (111.0) |

Figure 1A shows that substituters were more likely to be employed. The most common disorders and diseases were chronic pain, arthritis, anxiety, hypertension, cancer, migraines, depression, a sleep disorder, asthma, and Irritable Bowel Syndrome. Substituters had 25.4% more health conditions and diseases than non-substituters (Figure 1B).

**Figure 1.**
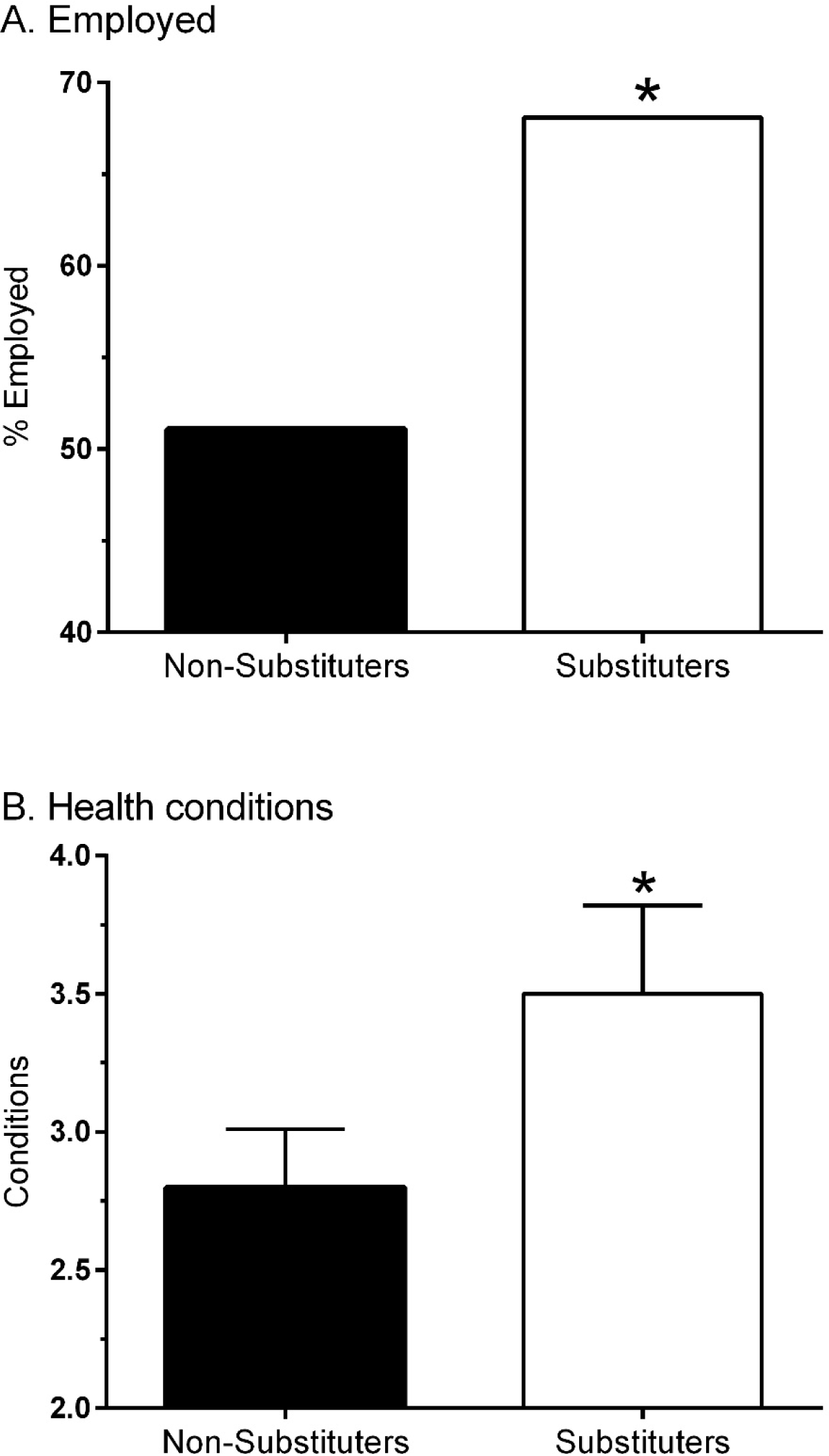
Employment (A) and total number (± SEM) of health conditions and diseases (B) among patients that did, or did not, reduce their alcohol consumption after starting Medical Cannabis. * *p* < .05.

## DISCUSSION

This study documents the self-reported efficacy of MC as a substitute for alcohol. As noted previously (Piper et al. 2017a), over two-fifths of dispensary members who used alcohol reported a reduction in alcohol use after starting MC. This is consistent with previous studies that documented a similar, or greater, reduction in alcohol (Reiman, 2006; Lucas et al. 2015). In a sample from California where the average age was a decade younger and half currently drank alcohol, forty percent used cannabis as a substitute for alcohol (Reiman, 2009). The current sample was composed primarily of chronic pain patients and not individuals who were especially motivated to make a change in their alcohol intake. It is an empirical question whether these results would generalize to others. Unfortunately, as existing medications are ineffective for the preponderance of alcoholics (Jonas et al. 2014), there is a tremendous need for novel pharmacotherapies, even if they only benefit a subset of patients, which can be employed with evidence based psychosocial treatments (Martin & Rehm, 2012).

There are two mechanisms by which MC could substitute for alcohol. First, rats that self-administered alcohol increased the endocannabinoid 2-arachidonoylglycerol in the nucleus accumbens (Caille et al. 2011) indicating that alcohol and THC have some shared neural substrates. Although efforts to employ the cannabinoid (CB_1_) inverse agonist rimonabant to decrease alcohol consumption were unsuccessful (George et al. 2010; Soyka et al. 2008), these findings should be interpreted with caution as rimonabant has a nonselective mechanism (Seeley et al. 2012) and may cause dysphoria. Second, MC is not covered by insurance and is expensive (>3,000 $USD/year, Piper et al. 2017b). However, we view a pharmacoeconomic explanation, i.e. participants spend money that previously was used for alcohol on MC, as unlikely because alcohol substituters were significantly more likely to be employed than non-substituters.

Studies with rodents have generally found that administration of cannabinoid receptor antagonists decreases alcohol consumption while cannabinoid agonists cause the opposite effect (Sloan et al. 2017). However, cannabidiol administration decreases alcohol consumption in mice (Viudez-Martínez et al. 2017) and blocks the neurotoxic effects of an alcohol binge (Hamelink, Hampson, Wink, et al. 2005; Liput, Hammell, & Stinchcomb, et al. 2013). Therefore, it is important to emphasize that MC is not a single homogenous product. There were 1,987 “strains” listed in a cannabis database and the ones available for medical use were dispensary specific (Piper, 2018). MC includes both THC, a CB_1_ and CB_2_ partial agonist and CBD. Cannabidiol has a complex pharmacology including acting as an indirect CB_1_ and CB_2_ antagonist (Mechoulam, Peters, Murillo-Rodriguez, et al. 2007), which could counteract the effects of THC. The mechanism of THC is also complex including acting as a serotonin (5-HT_1A_) partial agonist (Russo, Burnett, Hall, et al. 2005), a G-protein coupled receptor 55 antagonist, stimulating neurogenesis in the hippocampus, and inhibingthe adenosine transporter (Campos, Moreira, Gomes, et al. 2012; Gaston & Friedman, 2017). Perhaps some of the individual differences in MC associated outcomes may be due to the tremendous variability in MC products. It is an open question whether the findings from investigations with recreational marijuana use and alcohol, although subtle and contradictory (Metrik, Spillane, Leventhal, et al. 2011; Subbaraman, Metrik, Patterson, et al. 2017), generalize to MC because recreational and medical users may differ on demographic and motivational factors. Further, the strains used by MC patients (Piper, 2018) may show only partial overlap with those used for recreational purposes.

Substituters reported that they have been diagnosed with more diseases and disorders than non-substituters. Given the higher rates of employment among substituters, it is likely that these fit a profile of social drinkers. It is possible that the greater number of conditions, both psychiatric and non-psychiatric, might contribute to a heightened motivation to make lifestyle modifications to improve their quality of life.

Despite controversial findings regarding their limited effectiveness (Ferri et al. 2006), AA and other twelve step programs are inexpensive to operate and ubiquitous in the US. AA is not supportive of pharmacotherapies being employed to treat alcoholism (Alcoholics Anonymous, 2011). This perspective may stigmatize patients that choose these therapies, form barriers to the implementation of evidence based medicine (Mattick, Breen, Kimber, et al. 2009; Nielsen, Larance, Degenhardt, et al. 2016) and may function as an impediment to the successful treatment of addiction. Although the literature for MC substitution effects (Bachhuber et al. 2014; Boehnke, Litinas, & Clauw, 2016; Bradford & Bradford, 2016, 2017; Corroon, Mischley, Sexton, 2017; Lucas et al. 2015; Mikuriya, 1970, 2004; Reiman 2006, 2009; Piper et al. 2017a) does not yet include prospective investigations or randomized controlled trials, we are cautiously optimistic that future research of MC substitution for alcohol or opioids will include these more rigorous designs which will, perhaps, support the view that addiction is a biopsychosocial disorder where pharmacological interventions that target the endocannabinoid neurotransmitter system (Sloan et al. 2017) are a component of multimodal treatments.

There are some limitations to this study. First, although the response rate to the survey was generally good (Maine = 47%, Vermont = 41%, Piper et al. 2017b), the number of respondents that reported regular alcohol use was on the low end relative to what might be anticipated based on national data (Grant et al. 2017) which could reflect that dispensary members are not-representative of the general population. Second, recall of consumption of alcohol or other drugs that acutely impair memory is recognized as imperfect and future research could be conducted in a laboratory environment (Penetar et al. 2015) or employ more objective measures (e.g. medical records). Third, although this was an exploratory study exploring a novel topic, the variables identified were not hypothesized a priori and should be interpreted with caution.

## CONCLUSION

This retrospective study identified some demographic and health variables that differentiated persons that substituted MC for alcohol from those that did not. Given the need for more efficacious AUD pharmacotherapies, additional prospective or controlled research of the alcohol substitution effect following MC with a sample with more advanced alcohol misuse patterns may be warranted.

## Data Availability

The online survey is included as a supplemental material. Our IRB approved consent precludes sharing of the raw data.

## ACKNOWLEDGEMENTS

Rebecca M. DeKeuster, MEd and Alexander T. Abess, MD aided in survey construction. Olapeju Simoyan, MD and Stephanie D Nichols, PharmD provided feedback on an earlier version of this manuscript.

